# Detection of Air and Surface Contamination by Severe Acute Respiratory Syndrome Coronavirus 2 (SARS-CoV-2) in Hospital Rooms of Infected Patients

**DOI:** 10.1101/2020.03.29.20046557

**Authors:** Po Ying Chia, Kristen Kelli Coleman, Yian Kim Tan, Sean Wei Xiang Ong, Marcus Gum, Sok Kiang Lau, Stephanie Sutjipto, Pei Hua Lee, Than The Son, Barnaby Edward Young, Donald K. Milton, Gregory C. Gray, Stephan Schuster, Timothy Barkham, Partha Prathim De, Shawn Vasoo, Monica Chan, Brenda Sze Peng Ang, Boon Huan Tan, Yee-Sin Leo, Oon-Tek Ng, Michelle Su Yen Wong, Kalisvar Marimuthu

## Abstract

Understanding the particle size distribution in the air and patterns of environmental contamination of SARS-CoV-2 is essential for infection prevention policies. We aimed to detect SARS-CoV-2 surface and air contamination and study associated patient-level factors. 245 surface samples were collected from 30 airborne infection isolation rooms of COVID-19 patients, and air sampling was conducted in 3 rooms.

Air sampling detected SARS-CoV-2 PCR-positive particles of sizes >4 µm and 1-4 µm in two rooms, which warrants further study of the airborne transmission potential of SARS-CoV-2. 56.7% of rooms had at least one environmental surface contaminated. High touch surface contamination was shown in ten (66.7%) out of 15 patients in the first week of illness, and three (20%) beyond the first week of illness (p = 0.010).

## Introduction

Severe acute respiratory syndrome coronavirus 2 (SARS-CoV-2) causing coronavirus disease 2019 (COVID-19) has spread globally and many countries are experiencing ongoing local transmission despite varying levels of control efforts. Understanding the different transmission routes of SARS-CoV-2 is crucial in planning effective interventions to break the chain of transmission. Although extensive surface contamination with SARS-CoV-2 by a symptomatic patient has been demonstrated ^1^, little is known about airborne transmission of SARS-CoV-2. It is also unknown if asymptomatic individuals pose the same environmental contamination risk as symptomatic ones, although viral shedding has been demonstrated to continue even after clinical recovery of COVID-19 patients ^2^. There are multiple reports of asymptomatic patients testing positive for SARS-CoV-2 ^3,4^, and the potential transmission of the virus by an asymptomatic person has been described ^5^. Therefore, viral contamination of the air and surfaces surrounding asymptomatic or recovering COVID-19 patients could have serious implications for outbreak control strategies. This knowledge gap is recognized in the Report of the WHO-China Joint Mission on Coronavirus 2019 ^6^.

The primary objective of our study was to identify potential patient-level risk factors for environmental contamination by SARS-CoV-2 by sampling the air and surfaces surrounding hospitalized COVID-19 patients at different stages of illness.

## Methods

### Study design, patient selection and data collection

We conducted this cross-sectional study in airborne infection isolation rooms (AIIRs) at the National Centre for Infectious Diseases, Singapore. These rooms had 12 air changes per hour, an average temperature of 23°C, relative humidity of 53 – 59%, and exhaust flow of 579.6 m^3^/h.

Patients with a SARS-CoV-2 infection confirmed by a polymerase chain reaction (PCR)- positive respiratory sample within the prior 72 hours were included. Clinical characteristics, including the presence of symptoms, day of illness, day of stay in the room, supplemental oxygen requirement, and baseline characteristics, were collected. One patient from a previously published pilot study on environmental sampling in the same facility (Patient 30; Supplemental Table 1) was also included in the current analysis ^1^.

### Air sampling

Six NIOSH BC 251 bioaerosol samplers were placed in each of three AIIRs in the general ward to collect air samples. Particles collected with the NIOSH sampler are distributed into three size fractions. Particles >4 μm in diameter are collected in a 15 mL centrifuge tube, particles 1-4 μm in diameter are collected in a 1.5 mL centrifuge tube, and particles <1 μm in diameter are collected in a self-assembled filter cassette containing a 37-mm diameter, polytetrafluoroethylene (PTFE) filter with 3μm pores. All NIOSH samplers were connected to either SKC AirCheck TOUCH Pumps or SKC Universal air sampling pumps set at a flow-rate of 3.5 L/min and run for four hours, collecting a total of 5,040 L of air from each patient’s room.

In the room of Patient 1, three NIOSH samplers were attached to each of two tripod stands and situated at different heights from the ground (1.2m, 0.9m, and 0.7m) near the air exhaust to capture particles from the unidirectional airflow in the room. Throughout the four-hour sampling period, Patient 1 was intermittently facing the NIOSH samplers while seated one meter from the first tripod and 2.1 meters from the second tripod. Four SKC 37mm PTFE filter (0.3μm pore size) cassettes were also distributed throughout the room and connected to SKC Universal air sampling pumps set at a flow-rate of 5 L/min, each collecting an additional 1,200 L of air from the room.

In the rooms of Patients 2 and 3, three NIOSH samplers were attached to each of two tripod stands and situated at different heights from the ground (1.2m, 0.9m, and 0.7m). Throughout the four-hour sampling period, Patients 2 and 3 remained in bed within 1 meter from all 6 NIOSH samplers (Supplementary Figure 1). Patient 3 was also talking on the phone for a significant proportion of time during sampling. Additional SKC pumps with PTFE filter cassettes were not used in the rooms of Patient 2 and 3.

The 6 NIOSH samples from each room were pooled prior to analysis, but the particle size fractions remained separated. Each sample pool was representative of 5,040 L air.

### Surface sampling

Surface samples were collected with Puritan® EnviroMax Plus pre-moistened macrofoam sterile swabs (25-88060). Eight to 20 surface samples were collected from each room. Five surfaces were designated high-touch surfaces, including the cardiac table, entire length of the bed rails including bed control panel and call bell, bedside locker, electrical switches on top of the beds, and chair in general ward rooms (Supplemental Figure 1). In ICU rooms, the ventilator and infusion pumps were sampled instead of the electrical switches on top of the beds and chair (Supplemental Figure 2). Air exhaust outlets and glass window surfaces were sampled in five rooms, including the three rooms in which air sampling was performed. Toilet seat and automatic flush button (one combined swab) were sampled in AIIR rooms in the general ward.

### Sample transfer and processing

All samples were immediately stored at 4°C in the hospital prior to transfer to a BSL-3 laboratory where samples were immediately processed and stored at -80°C unless directly analyzed. Prior to RNA extraction, NIOSH aerosol sample tubes and filters were processed as previously described ^7^, with slight modification due to the pooling of samples.

### Laboratory methods

The QIAamp viral RNA mini kit (Qiagen Hilden, Germany) was used for sample RNA extraction. Real-time PCR assays targeting the envelope (E) genes ^8^ and an in house orf1ab assay were used to detect SARS-CoV-2 in the samples ^9^. All samples were run in duplicate and with both assays. Positive detection was recorded as long as amplification was observed in at least 1 assay.

### Statistical analysis

Statistical analysis was performed using Stata version 15.1 (StataCorp, College Station, Texas) and GraphPad Prism 8.0 (GraphPad Software, Inc., San Diego). *P* <0.05 was considered statistically significant, and all tests were 2-tailed. For the surface environment, outcome measures analyzed were any positivity by room and pooled percentage positivity by day of illness and respiratory viral load (represented by clinical cycle threshold (Ct) value). We analyzed the factors associated with environmental contamination using the Student t-test, or the nonparametric Wilcoxon rank-sum test was used for continuous variables depending on their distribution. The χ2 or Fisher exact test was used to compare categorical variables. We plotted the best fit curve by least-square method to study the environmental contamination distribution across various the days of illness and clinical Ct value.

## Results

Environmental sampling was conducted in three AIIRs in the ICU and 27 AIIRs in the general ward. Air sampling was performed in three of the 27 AIIRs in the general ward. All patients reported COVID-19 symptoms. Seven patients (23%) were asymptomatic at the time of environmental sampling. Of the 23 symptomatic patients, 18 (78%) had respiratory symptoms, one had gastrointestinal symptoms, one had both respiratory and gastrointestinal symptoms, and three patients (10%) had fever or myalgia only (Supplemental Table 1).

**Table 1.** Severe acute respiratory syndrome coronavirus 2 (SARS-CoV-2) detections in the air of hospital rooms of infected patients

| Patient | Day of illness | Symptoms reported on day of air sampling | Clinical Ct value* | Airborne SARS-CoV-2 concentrations (RNA copies m<sup>-3</sup> air) | Aerosol particle size | Samplers used |
| --- | --- | --- | --- | --- | --- | --- |
| 1 | 9 | Cough, nausea, dyspnea | 33.22 | ND | -- | NIOSH |
|  |  |  |  | ND | -- | SKC Filters |
| 2 | 5 | Cough, dyspnea | 18.45 | 2,000 | >4 µm | NIOSH |
|  |  |  |  | 1,384 | 1-4 µm |  |
| 3 | 5 | Asymptomatic<sup>†</sup> | 20.11 | 927 | >4 µm | NIOSH |
|  |  |  |  | 916 | 1-4 µm |  |

ND = none detected
\*PCR cycle threshold value from patient's clinical sample
<sup>†</sup>Patient reported fever, cough, and sore throat until the day before the sampling. Patient reported no symptoms on the day of sampling, however was observed to be coughing during sampling

Air samples from two (66.7%) of three AIIRs tested positive for SARS-CoV-2, in particle sizes >4 µm and 1-4 µm in diameter (Table 1). Total SARS-CoV-2 concentrations in air ranged from 1.84×10^3^ to 3.38×10^3^ RNA copies per m^3^ air sampled. Rooms with viral particles detected in the air also had surface contamination detected.

There were no baseline differences between patients with environmental surface contamination and those without, in terms of age, comorbidities, and positive clinical sample on the day of sampling. Median cycle threshold (Ct) values of the clinical specimens for patients with and without environmental surface contamination were 25.69 (IQR 20.37 to 34.48) and 33.04 (28.45 to 35.66) respectively (Table 2).

**Table 2:** Baseline clinical characteristics of COVID-19 patients with environmental contamination

| Characteristics of COVID-19 patients | Rooms with surface environment contamination (n=17) | Rooms without surface environment contamination (n=13) | P-value |
| --- | --- | --- | --- |
| Median age (IQR) | 52 (42 to 62) | 44 (36 to 55) | 0.75 |
| Male Sex (%) | 6 (46%) | 8 (47%) | 0.96 |
| Median Age Adjusted Charlson's Comorbidity Index (IQR) | 1 (0 to 2) | 1 (0 to 1) | 0.69 |
| Median day of Illness (IQR) | 5 (4 to 9) | 13 (5 to 20) | 0.17 |
| Median day of stay in room (IQR) | 3 (3 to 8) | 4 (2 to 16) | 0.95 |
| Oxygen requirement (%) | 0 | 4 (31) | 0.03 |
| Symptomatic (%) | 12 (71) | 11 (85) | 0.43 |
| Respiratory symptoms (%) | 11 (65) | 7 (54) | 0.55 |
| Gastrointestinal symptoms (%) | 1 (6) | 1 (8) | >0.99 |
| Clinical Cycle threshold value, median (IQR)* | 25.69 (20.37 to 34.48) | 33.04 (28.45 to 35.66) | 0.06 |

\*PCR cycle threshold value from patient's clinical sample

Of the rooms with environmental contamination, the floor was most likely to be contaminated (65%), followed by the bed rail (59%), and bedside locker (42%) (Figure 1). Contamination of toilet seat and automatic toilet flush button was detected in five out of 27 rooms, and all five occupants had reported gastrointestinal symptoms within the preceding one week of sampling. We did not detect surface contamination in any of the three ICU rooms.

**Figure 1:**
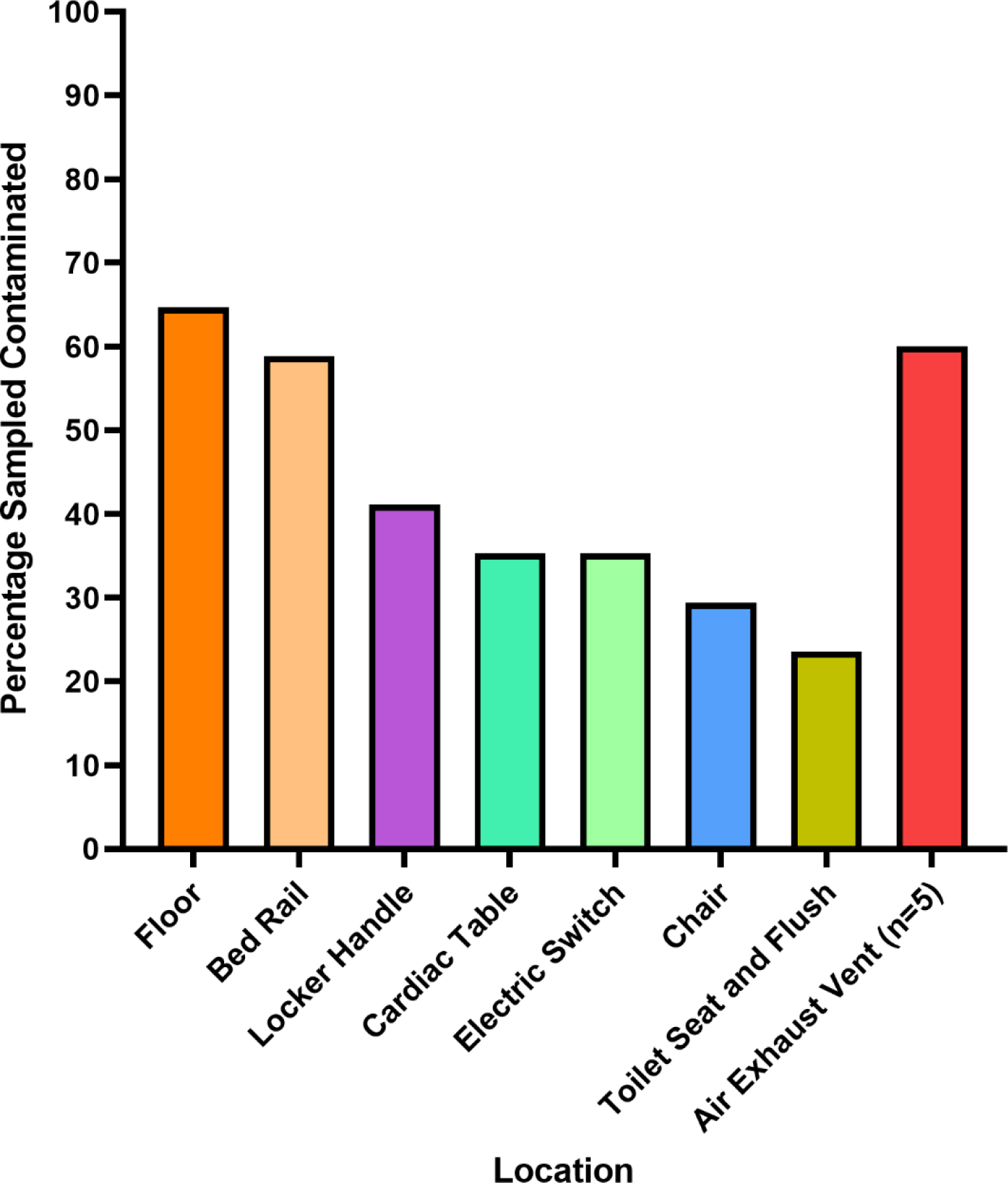
Percentage of contaminated swabs from surface samples, in rooms with any contamination. All other sites were n=17, except for air exhaust vents where n=5

Presence of environmental surface contamination was higher in week 1 of illness (Figure 2) and showed association with the clinical cyclical threshold (P=0.06). Surface environment contamination was not associated with the presence of symptoms or supplementary oxygen (Table 2). In a subgroup analysis, the presence and extent of high-touch surface contamination were significantly higher in rooms of patients in their first week of illness (Figure 2). The best fit curve with the least-squares fit (Figure 3) showed that the extent of high-touch surface contamination declined with increasing duration of illness and Ct values. There was also no correlation between the Ct values of clinical samples and the Ct values of environmental samples across the days of illness (Supplemental Figure 3).

**Figure 2:**
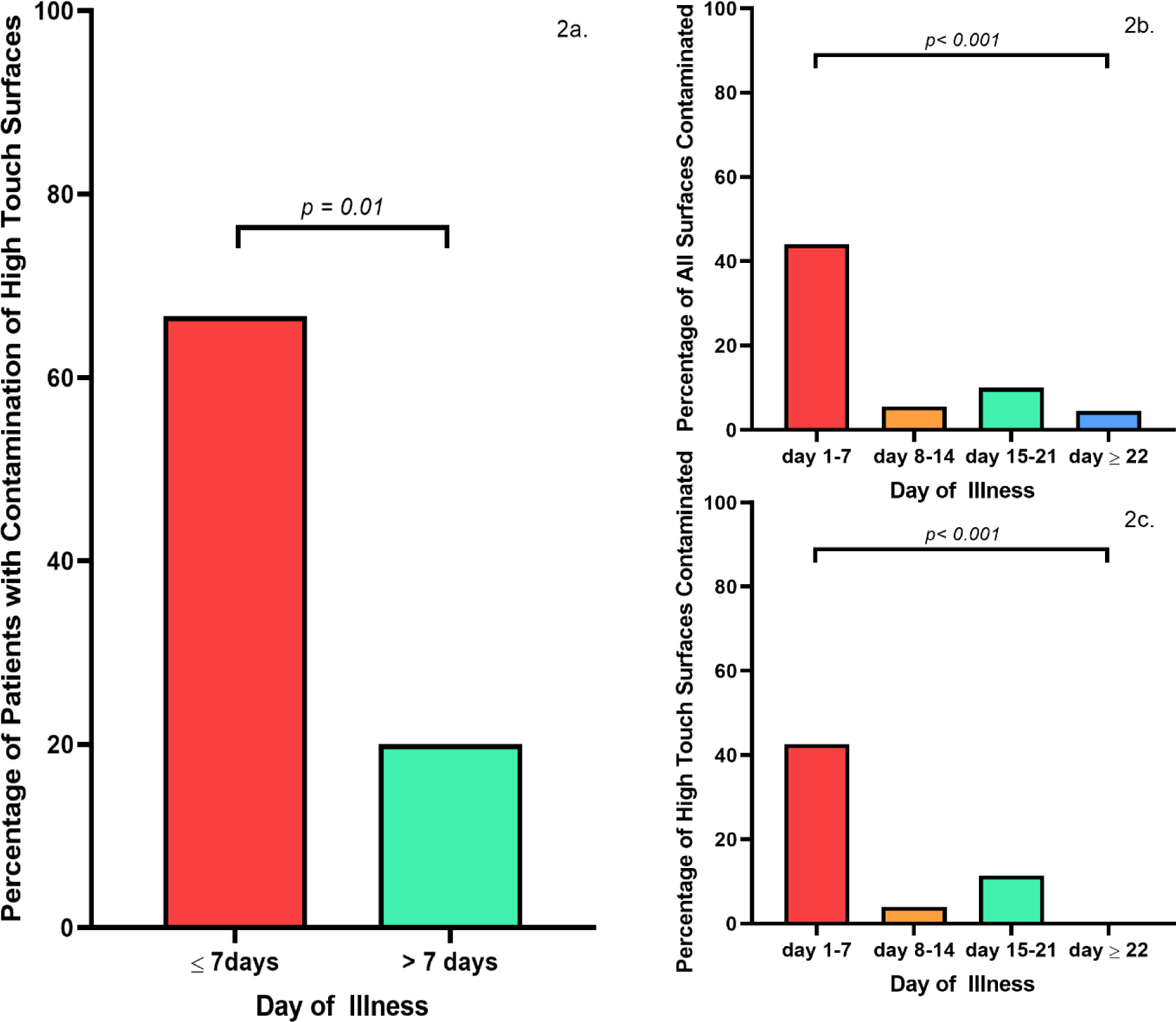
2a. Percentage of patients with contamination of high touch surfaces in in the first week of illness compared with more than first week of illness. 2b. Percentage of surfaces contaminated across weeks of illness. 2c. Percentage of high-touch surfaces contaminated across weeks of illness

**Figure 3:**
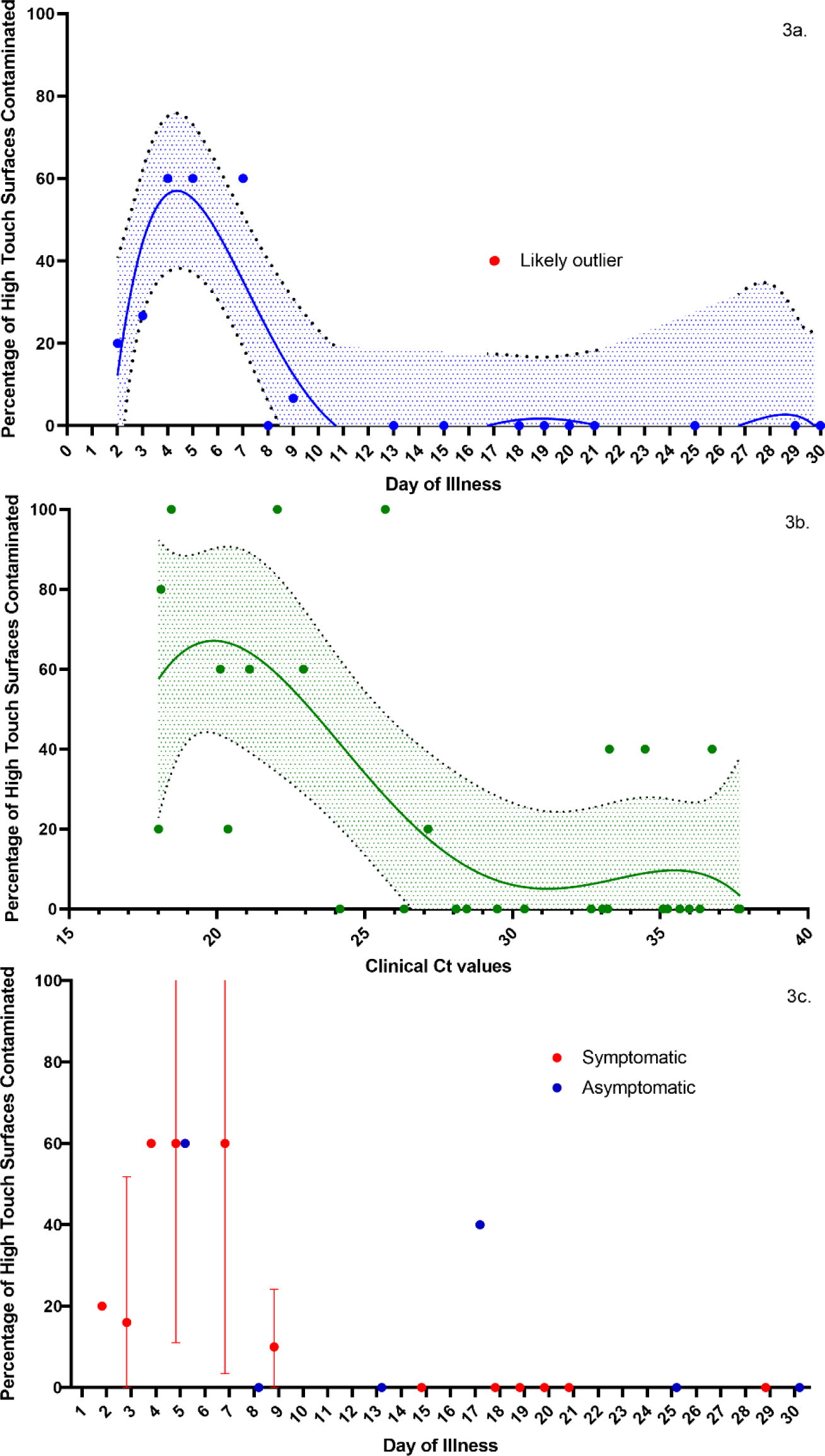
3a. Mean percentage of high touch surface contaminated by day of illness with 95% confidence interval with best fit curve. 3b. Mean percentage of high touch surfaces contaminated by clinical cycle threshold values with 955 confidence interval with best fit curve. 3c._Mean percentage of high touch surface contaminated by day of illness with 95% confidence interval grouped by symptoms

## Discussion

Surface sampling revealed that the PCR-positivity high-touch surfaces was associated with nasopharyngeal viral loads and peaked at approximately day four to five of symptoms. Air sampling of the AIIR environments of two COVID-19 patients (both day five of illness with high nasopharyngeal swab viral loads) detected the presence of SARS-CoV-2 particles sized 1-4 µm and > 4 µm. The absence of any detection of SARS-CoV-2 in air samples of the third patient (day nine of illness with lower nasopharyngeal viral load concentration) suggests that the presence of SARS-CoV-2 in the air is possibly highest in the first week of illness.

Recent aggregated environmental sampling and laboratory experiments have examined the particle size distribution of SARS-CoV-2 in the air. A study from Wuhan, China sampled three different environmental settings and detected aerosol size range particles ^10^.

Additionally, a recent laboratory study demonstrated the ability of SARS-CoV-2 to remain viable in aerosols for up to 3 hours ^11^. While limited in subject numbers, our study examined this issue at the individual patient-level, thus enabling correlation of particle size distribution in the air with symptoms duration and nasopharyngeal viral loads. The absence of aerosol-generating procedures or intranasal oxygen supplementation reduces the possibility of our current findings being iatrogenic in nature. Larger individual patient-level studies examining the droplet and aerosolizing potential of SARS-CoV-2 over different distances and under different patient and environmental conditions are rapidly needed to determine the generalizability of our current findings.

In the current analysis the presence and concentration of SARS-CoV-2 in air and high-touch surface samples correlated with the day of illness and nasopharyngeal viral loads of COVID-19 patients. This finding is supported by multiple observational clinical studies have demonstrated that SARS-CoV-2 viral loads peak in the first week among COVID-19 patients ^2,12,13^, with active viral replication in the upper respiratory tract in the first five days of illness^14^. This finding could help inform public health and infection prevention measures in prioritizing resources by risk stratifying COVID-19 patients by their potential to directly or indirectly transmit the SARS-CoV-2 virus to others.

Our study was limited in that it did not determine the ability of SARS-CoV-2 to be cultured from the environmental swabs and the differentially-sized air particles which would be vital to determining the infectiousness of the detected particles. Another study from Nebraska attempted virus culture on SARS-CoV-2 PCR-positive air samples, however could not isolate viable virus ^15^. The difficulty in culturing virus from air samples arises from low virus concentrations, as well as the compromised integrity of the virus due to air sampling stressors. Future studies using enhanced virus culture techniques could be considered ^16^, and efforts to design a culture method to isolate virus from our samples is underway. Second, sampling in an AIIR environment may not be representative of community settings and further work is needed to generalize our current findings. Third, we sampled each room at a single timepoint during the course of illness and did not track environmental contamination over the course of illness for individual patients. Fourth, as clinical results were within 72 hours of environmental testing, it is plausible that during the day of testing, viral load was actually low or negligible, hence limiting environmental contamination.

Current evidence does not seem to point to aerosolization as the key route of transmission of SARS-CoV-2, and there have been reports of healthcare workers not being infected after exposure to confirmed patients despite not using airborne precautions^17^. Detailed epidemiologic studies of outbreaks, in both healthcare and non-healthcare settings, should be carried out to determine the relative contribution of various routes of transmission and their correlation with patient-level factors.

In conclusion, in a limited number of AIIR environments, our current study involving individual COVID-19 patients not undergoing aerosol-generating procedures or oxygen supplementation suggest that SARS-CoV-2 can be shed in the air from a patient in particles sized between 1 to 4 microns. Even though particles in this size range have the potential to linger longer in the air, more data on viability and infectiousness of the virus would be required to confirm the potential airborne spread of SARS-CoV-2. Additionally, the concentrations of SARS-CoV-2 in the air and high-touch surfaces could be highest during the first week of COVID-19 illness. Further work is urgently needed to examine these findings in larger numbers and different settings to better understand the factors affecting air and surface spread of SARS-CoV-2 and inform effective infection prevention policies.

## Data Availability

All data referred to in the manuscript has been stored on a secure server with access by the authors.

## Acknowledgements

We thank Dr Bill Lindsley (National Institute for Occupational Safety and Health, NIOSH) for loaning us aerosol samplers and guiding us in their use; Dr. Raquel Binder and Emily Robie from the Duke One Health team for quickly providing us equipment for use in this study; Dr Hsu Li Yang (Saw Swee Hock School of Public Health, National University of Singapore) for facilitating the air sampling; and Dr Ding Ying (National Centre for Infectious Diseases, Singapore) for helping with project coordination. No compensation was received for their roles in the study.

We further thank the colleagues in DSO National Laboratories in the environmental detection team and clinical diagnostics team for BSL3 sample processing and analysis as well as the logistics and repository team for transport of biohazard material, inventory and safekeeping of received items.

